# Access to reproductive health services: findings from the 2023 Reproductive Health Survey for England

**DOI:** 10.64898/2026.06.26.26356647

**Authors:** Ona L McCarthy, Melissa J Palmer, Cecile Knai, Emily Warren, Beth Jakubowski, Agata Pacho, Rebecca S French

## Abstract

**Background:** Recent research has documented poor reproductive health among women and people assigned female in England. Access to reproductive health services is hindered by an opaque and fragmented system.

**Methods:** We conducted the 2023 Reproductive Health Survey for England, a non-probability online survey, in September and October 2023 (N = 59,332). In this analysis, we examined access to reproductive health services across three domains: heavy menstrual bleeding and severe pain, gynaecological symptoms and conditions and menopause-related symptoms. Weighting the sample to match the 2021 Census age distribution, we assessed differences by ethnic group, subjective financial situation, educational attainment and region across the domains using logistic regression analysis and controlling for age.

**Results:** Respondents reported low access to reproductive health services overall, including 34.8% (8,644/24,952) of those with heavy bleeding or severe period pain, 44.7% (6,709/15,569) with menopausal symptoms and 55.3% (21,010/37,411) with gynaecological symptoms or conditions. When controlling for age, there were decreased odds of service access for menopause-related symptoms and increased odds of service access for gynaecological symptoms or conditions among Black ethnic groups. Respondents with a higher education degree had greater access to services for heavy bleeding or severe pain and gynaecological symptoms and conditions. Compared to London, all other regions had lower access to services for heavy bleeding or severe pain. Satisfaction ranged from 16.5% (741/4,666) for polycystic ovary syndrome services to 80.2% (166/207) for reproductive cancer services.

**Conclusions:** Access to reproductive health services is poor in England and requires urgent action to address barriers to access.

## Background

Equitable access to high-quality reproductive health services has been on the global agenda for several decades now. The 2025 report of a United Nations special programme on human reproduction notes that while great progress has been made in research and action around contraception, abortion laws, maternal mortality and other key areas, considerable inequalities and inconsistencies in access to essential reproductive health services remain at global level (1). High-income countries are no exception with regard to persistent reproductive health inequalities (2). In England, levels of reproductive health burden amongst women and people assigned female at birth (hereafter referred to as women) aged 16-55 are high, with nearly three-quarters (74%) experiencing reproductive symptoms or conditions over the past year (3). Twenty-eight percent of survey respondents had experienced at least one reproductive morbidity, with menstrual-related morbidities (severe period pain, heavy bleeding or hot flushes or night sweats) the most prevalent, at 62%. Recent studies indicate that Asian and Black women and women who live in the most deprived areas of England have higher rates of maternal (4) and perinatal mortality (5), compared to White women and women living in the least deprived areas. There is an urgent need to understand the structural drivers of these reproductive health experiences and outcomes.

In England, a range of services provide reproductive health care, including primary care, community sexual and reproductive health (SRH) clinics, pharmacies, and gynaecology services. However, fragmented commissioning of services still present barriers in identifying and accessing appropriate services (6). Providing better integrated care can ensure reproductive health needs are addressed together, such as being able to have a cervical smear and intrauterine device insertion done in one visit rather than two visits to separate services (7). Lack of integrated care is not just a personal inconvenience, but it can result in delays in diagnosis and treatment, and financial costs for patients and the NHS (6). An opaque and difficult to navigate system can also widen existing reproductive health inequities and put underserved groups at a further disadvantage. Past experiences and expectations of being dismissed by healthcare professionals when reporting symptoms may influence decisions to access healthcare services and ultimately lead to long delays in obtaining diagnosis, treatment and support (8, 9).

Further compounding these problems is the recently reported lack of awareness and understanding of women’s reproductive health conditions (such as endometriosis, adenomyosis and heavy menstrual bleeding) and beliefs among some health professionals that people-particularly from minority ethnic groups-exaggerate the severity of their experiences (6). The government report recommends that medical misogyny and racism in reproductive health care be addressed by primary care training programmes to challenge racial biases and to not dismiss or normalise patient reports of pain. Recent policy positions regarding ethnicity and deprivation in women’s health by the Royal College of Obstetricians & Gynaecologists (RCOG) recommend urgent action to reduce inequalities by removing barriers to care (10, 11).

The 2022 England’s Women’s Health Strategy for England (12) set out a life-course approach to women’s health. One of the goals of the strategy was to improve women’s experiences of healthcare support. Women’s Health Hubs have been promoted as a means of ensuring a more holistic and joined up approach to women’s health, but they continue to be relatively small in number, and their impact has not been established (13). In 2025, the RCOG specifically highlight the need for “easy access comprehensive support for gynaecological conditions, menstrual health and menopause” and note that these areas should continue to be “a core function of a Women’s Health Strategy” given their predictability and that they are common features of women’s lives (14).

In this paper, we report on access to and satisfaction with reproductive health services using data collected from the 2023 Reproductive Health Survey for England (RHSE 2023) (3, 15). The overall aim of this analysis is to characterise access to reproductive health services in England, using data from the RHSE 2023. Specific research objectives are to: 1) identify potential need for health services amongst women and those assigned female at birth across the three specified reproductive health domains: heavy menstrual bleeding and severe pain, gynaecological symptoms and conditions and menopause-related symptoms; 2) estimate the percentage in the population who access services for their reproductive symptoms or conditions; 3) explore if key demographic characteristics are associated with services access among those with a need and 4) amongst those who have accessed services, the percent satisfied with the service.

## Methods

### Study design & participants

The RHSE 2023 was a non-probability online survey, conducted between 7 September and 19 October 2023 (3, 15). The survey recruited respondents through social media advertising, social media posts and network dissemination through our partner organisations (LGBT Foundation; Race Equality Foundation; Brook; and Birth Companions) and through survey launch press releases. People were eligible to complete the survey if they were assigned female at birth, aged 16–55 years, and resident in England. There total sample was 59,332 and compared to the 2021 Census, the sample over-represented people aged 20-34 and with higher education levels, and under-represented people from minority ethnic groups.

### Data

The data used in this analysis include demographic variables (age, ethnic group, subjective financial situation, educational attainment and region) and variables on access to and satisfaction with services across the three domains.

#### Heavy bleeding or severe period pain

Respondents who indicated that they needed to change their pad, tampon, or menstrual cup every 1-2 hours were characterised as having ‘heavy bleeding’. Participants who indicated that they had had a period in the last 12 months were asked how much pain they had experienced in general during their period in the last year (no pain, mild pain, moderate pain and severe pain). For this analysis we combined those who indicated that they experience severe pain and heavy bleeding into one variable, which indicated a service need. We estimated the population-level service need as the percentage of respondents experiencing the symptoms, with all respondents who answer the routing question in the denominator (includes those who have not had a period in over a year).

#### Gynaecological symptoms and conditions

All respondents were asked if they experienced gynaecological symptoms in the last year and currently experience conditions. The symptoms respondents were asked about were urinary incontinence; pelvic pain (not including period pain); pain during or after sex; vulval pain/problems; vaginal bleeding after sex and abnormal vaginal discharge. The conditions respondents were asked about were polycystic ovary syndrome (PCOS); endometriosis; uterine fibroids; uterine or cervical polyps; pelvic organ prolapse; cervical, ovarian, uterine or breast cancer. Respondents who reported any symptom or condition were identified as having a service need for gynaecological care.

#### Menopause-related symptoms

Respondents aged 40-55 were asked if they experienced symptoms that “may be related to perimenopause or menopause” in the last year. These symptoms were listed as: hot flushes; night sweats; vaginal dryness; discomfort during sex; reduced libido; difficulty sleeping; low mood or anxiety and problems with memory or concentration. Respondents who reported at least one of these symptoms were identified as having a service need for menopause-related care.

#### Access to services

Following each set of questions regarding need in each domain, respondents who indicated a need were asked if they received help or advice from a healthcare professional for their symptoms or conditions in the last year. For heavy bleeding or pain, this was one question with yes or no response options. For gynaecological symptoms and conditions, respondents who indicated a symptom or condition were asked if they accessed a variety of services for these in the past year. Respondents who indicated that they accessed a sexual health/family planning/reproductive health clinic, GP surgery, Women’s Health Hub, Hospital, private doctor or clinic were identified as having access a service. All respondents over 40 (regardless of whether they reported any menopause-related symptom) were asked if they received help or advice from a healthcare professional about menopause-related concerns in the last year, with yes or no response options. For this analysis, access to services was considered among respondents with a need.

#### Treatment

Respondents who reported having heavy bleeding were asked if they had treatment specifically for their heavy bleeding in the last year. Respondents could select multiple responses from the following treatment options: hormonal tablet, hormonal IUS, hormonal injection, hormonal implant (contraceptive implant, non-hormonal medication (e.g. Tranexamic Acid), minor surgery (e.g. endometrial ablation, hysterectomy, other treatment). All respondents aged 40 and over were asked if they used treatment to help with menopause-related symptoms in the last year, regardless of whether or not they indicated any symptoms. Respondents could select multiple responses and were considered as having received treatment if they chose at least one from the following treatment options: hormone replacement therapy, testosterone, hormonal IUS, other hormonal pharmaceutical treatments, anti-depressants and other medications.

#### Satisfaction

Respondents with heavy bleeding or period pain who indicated that they received help or advice from a healthcare professional about their symptoms in the past year were asked how satisfied they were with the care received (for this analysis we limited it to those who indicated heavy bleeding or *severe* period pain). Respondents were asked how satisfied they were with the care they received for each of gynaecological symptoms or conditions that they reported. Respondents aged 40 and over who indicated that they received help or advice from a healthcare professional about menopause-related concerns in the past year were asked how satisfied they were with the help or advice received. Satisfaction with help or advice for both of these domains was classified as satisfied/very satisfied or other (neither satisfied or dissatisfied, dissatisfied or very dissatisfied).

### Statistical analysis

For the service access analyses we weighted the sample to match the 2021 Census age distribution due to the over-representation of respondents in the 20-34 age group. We assessed differences in access by ethnic group, subjective financial situation, educational attainment and region across the domains using logistic regression, controlling for age. Statistical significance was considered at p<0.05 for all analyses. StataNow/SE 18.5 was used for all analyses.

## Results

### Respondent characteristics

Missing data on key variables was around 4% for each, with one at 11.5% (gynaecological symptoms and conditions).

Table 1 reports the respondent characteristics of the unweighted sample. The large majority of respondents identified as woman/girl (data not shown in Table 1, 96.2%, 56,822/59,082). Compared to 2021 Census data, this sample has an over-representation of White ethnic groups (92.3% vs 77.9% in the Census) and an over-representation of people with a degree or equivalent (72.6% vs 41.4% in the Census) (16).

**Table 1.** Respondent characteristics*.

|  | % | 95% CI | n | 2021 Census |
| --- | --- | --- | --- | --- |
| **Age group** |  |  | **N=59,332** |  |
| 16 to 19 | 6.7 | (6.53-6.94) | 3,995 | 8.4% |
| 20 to 24 | 15.4 | (15.15-15.73) | 9,159 | 11.4% |
| 25 to 29 | 16.7 | (16.44-17.04) | 9,931 | 12.7% |
| 30 to 34 | 16.7 | (16.39-16.99) | 9,904 | 13.7% |
| 35 to 39 | 13.4 | (13.13-13.68) | 7,954 | 13.1% |
| 40 to 44 | 11.3 | (11.05-11.56) | 6,708 | 12.3% |
| 45 to 49 | 8.7 | (8.45-8.90) | 5,147 | 12.3% |
| 50 to 55 | 11.0 | (10.76-11.27) | 6,534 | 16.0% |
| **Ethnicity census groupings** |  |  | **N= 58,181** |  |
| White | 92.3 | (92.06-92.49) | 53,687 | 77.9% |
| Mixed or Multiple ethnic groups | 3.3 | (3.18-3.47) | 1,931 | 2.9% |
| Asian or Asian British | 2.5 | (2.38-2.63) | 1,457 | 11.6% |
| Black, Black British, Caribbean or African | 1.4 | (1.28-1.46) | 795 | 5.1% |
| Another ethnic group | 0.5 | (0.48-0.60) | 311 | 2.6% |
| **How managing financially** |  |  | **N= 57,653** |  |
| Living comfortably | 21.5 | (21.18-21.85) | 12,405 |  |
| Doing alright | 44.5 | (44.11-44.92) | 25,662 |  |
| Just getting by | 23.5 | (23.13-23.82) | 13,533 |  |
| Finding it quite difficult | 7.8 | (7.63-8.07) | 4,522 |  |
| Finding it very difficult | 2.7 | (2.53-2.79) | 1,531 |  |
| **Has degree or equivalent** |  |  | **N=57,377** |  |
| No | 27.4 | (27.07-27.80) | 15,738 | 58.6% |
| Yes | 72.6 | (72.20-72.93) | 41,639 | 41.4% |
| **Government Region** |  |  | **N=52,476** |  |
| Northeast | 3.9 | (3.76-4.09) | 2,056 | 4.5% |
| Northwest | 12.1 | (11.79-12.35) | 6,333 | 12.9% |
| Yorkshire and Humber | 10.5 | (10.24-10.77) | 5,512 | 9.5% |
| East Midlands | 8.2 | (7.99-8.46) | 4,313 | 8.4% |
| West Midlands | 8.2 | (7.98-8.45) | 4,307 | 10.3% |
| East | 10.6 | (10.34-10.87) | 5,562 | 10.9% |
| London | 17.8 | (17.48-18.13) | 9,342 | 18.2% |
| Southeast | 16.6 | (16.24-16.87) | 8,687 | 16.0% |

|  | **%** | **95% CI** | **n** | **2021**<br>**Census** |
| --- | --- | --- | --- | --- |
| Southwest | 12.1 | (11.85-12.41) | 6,364 | 9.4% |

*\* Respondent characteristics based on the unweighted sample*

### Access to services

Table 2 reports services access among those with a need, by domain. Among respondents who had a period in the last year and either reported needing to change their pad, tampon, or menstrual cup every 1-2 hours or reported severe pain during their period (44.1%, 25,990/56,920), only 34.8% (8,644/24,952) said they received help or advice and 35.6% (8,009/22,388) said they received treatment (N.B. receiving help or advice and receiving treatment are not mutually exclusive). Despite almost all respondents aged 40-55 reporting at least one menopause-related symptom (91.9%, 15,642/17,157), less than half (44.7%, 6,709/15,569) received help or advice and less than half (47.7%, 7,166/15,569) received treatment for their symptoms. Compared to the other domains, access to services was slightly higher among respondents who reported any gynaecological symptom or condition (70.7%, 37,411/52,511), at 55.3% (21,010/37,411).

**Table 2.**
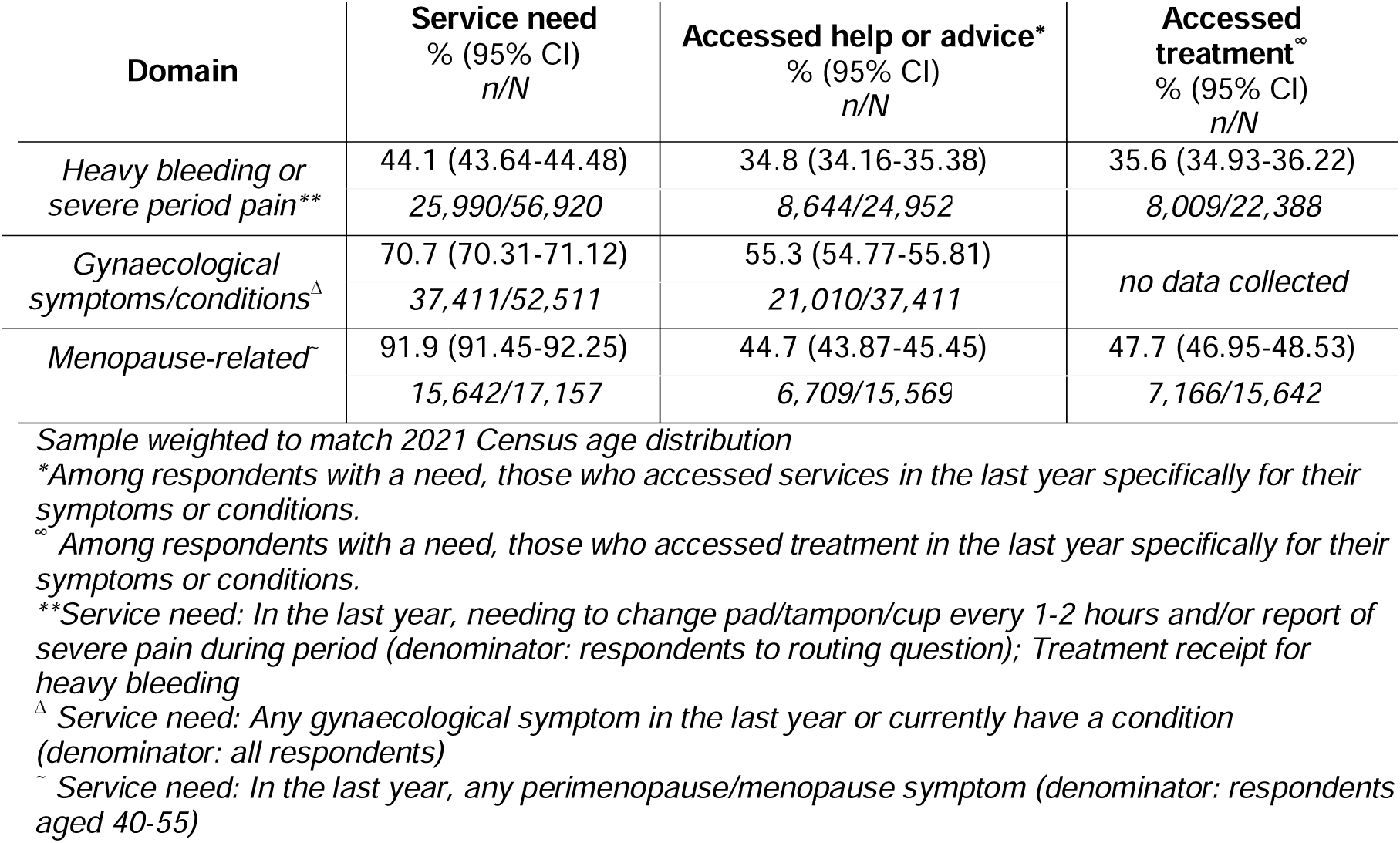
Access to services.

| Domain | Service need<br>% (95% CI)<br>n/N | Accessed help or advice*<br>% (95% CI)<br>n/N | Accessed treatment<sup>∞</sup><br>% (95% CI)<br>n/N |
| --- | --- | --- | --- |
| Heavy bleeding or severe period pain** | 44.1 (43.64-44.48)<br>25,990/56,920 | 34.8 (34.16-35.38)<br>8,644/24,952 | 35.6 (34.93-36.22)<br>8,009/22,388 |
| Gynaecological symptoms/conditions<sup>Δ</sup> | 70.7 (70.31-71.12)<br>37,411/52,511 | 55.3 (54.77-55.81)<br>21,010/37,411 | no data collected |
| Menopause-related<sup>ˆ</sup> | 91.9 (91.45-92.25)<br>15,642/17,157 | 44.7 (43.87-45.45)<br>6,709/15,569 | 47.7 (46.95-48.53)<br>7,166/15,642 |

Sample weighted to match 2021 Census age distribution
\*Among respondents with a need, those who accessed services in the last year specifically for their symptoms or conditions.
<sup>∞</sup> Among respondents with a need, those who accessed treatment in the last year specifically for their symptoms or conditions.
\*\*Service need: In the last year, needing to change pad/tampon/cup every 1-2 hours and/or report of severe pain during period (denominator: respondents to routing question); Treatment receipt for heavy bleeding
<sup>Δ</sup> Service need: Any gynaecological symptom in the last year or currently have a condition (denominator: all respondents)
<sup>ˆ</sup> Service need: In the last year, any perimenopause/menopause symptom (denominator: respondents aged 40-55)

Table 3 presents the percentages and age-adjusted associations between access to services and subjective financial situation, ethnicity, educational attainment and region. Compared to White groups, Black ethnic groups with a need for services had a higher percentage of access to services for gynaecological symptoms and conditions, at 63.5% (325/506) vs 55.0% (54.41-55.50). There was little variation in access to services by educational attainment. In terms of region, access to services for heavy bleeding or severe period pain and gynaecological symptoms and conditions was highest in London 56.7%, (856/1,483) and 57.5%, (3,371/5,791), respectively.

**Table 3.**
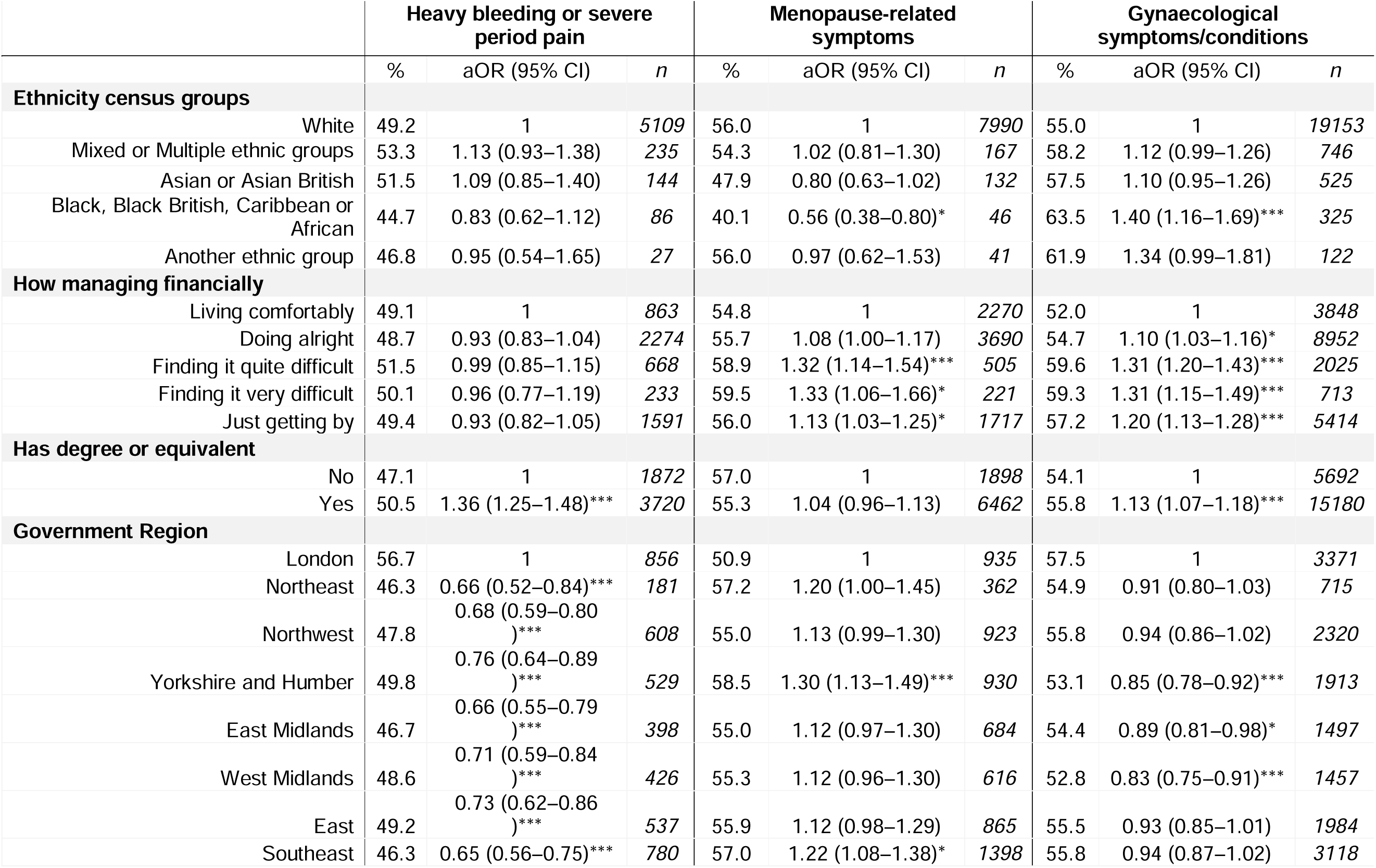
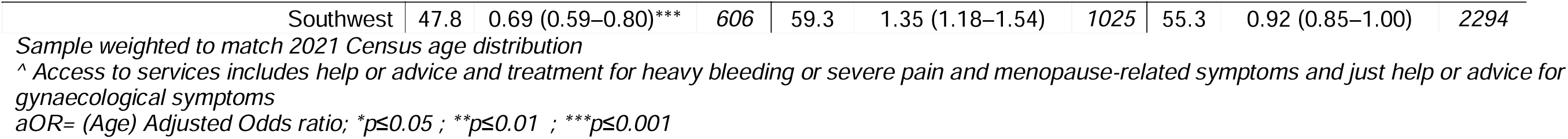
Association between demographic characteristics and access to services^.

Respondents finding it ‘quite’ (aOR 1.32, 95% CI 1.14–1.54) or ‘very’ (aOR 1.33, 95% CI 1.06–1.66) difficult or ‘just getting by’ (aOR 1.13, 95% CI 1.03–1.25) had greater odds of accessing services for menopause-related symptoms compared to those ‘living comfortably’. Compared to those living comfortably, all other groups had greater odds of accessing service for gynaecological symptoms or conditions. Respondents identifying as Black, Black British, Caribbean or African had almost half the odds (aOR 0.56, 95% CI 0.38–0.80) of accessing menopause-related services and 1.40 times the odds (95% CI 1.16–1.69) of accessing gynaecological services compared to respondents in the White group. Respondents with a degree had greater odds of accessing services for heavy bleeding or severe period pain (aOR1.36, 95% CI 1.25–1.48) and gynaecological symptoms and conditions (aOR 1.13, 95% CI 1.07–1.18). Compared to respondents living in London, respondents living in other England regions all had lower odds of accessing services for heavy bleeding or severe period pain. Compared to London, respondents living in Yorkshire and Humber (aOR 1.30, 95% CI 1.13–1.49) and the Southeast (aOR 1.22, 95% CI 1.08–1.38) had greater odds of accessing services for menopause-related symptoms. Regarding gynaecological symptoms and conditions, respondents living in Yorkshire and Humber (aOR 0.85, 95% CI 0.78–0.92), the East (aOR 0.89, 95% CI 0.85–1.01) and West Midlands (aOR 0.83, 95% CI 0.75–0.91) had decreased odds of accessing services.

Table 4 presents the percent satisfied with services accessed in the last year for symptoms and conditions in each domain. The percent satisfied varied from 16.5% (741/4,666) among those who accessed services for PCOS and up to 80.2% (166/207) among respondents who accessed cancer services. While satisfaction with the eight types of services varied, it was low overall, with the percent satisfied/very satisfied below 50% for all but cancer (80%) and menopause-related services (60%).

**Table 4.**
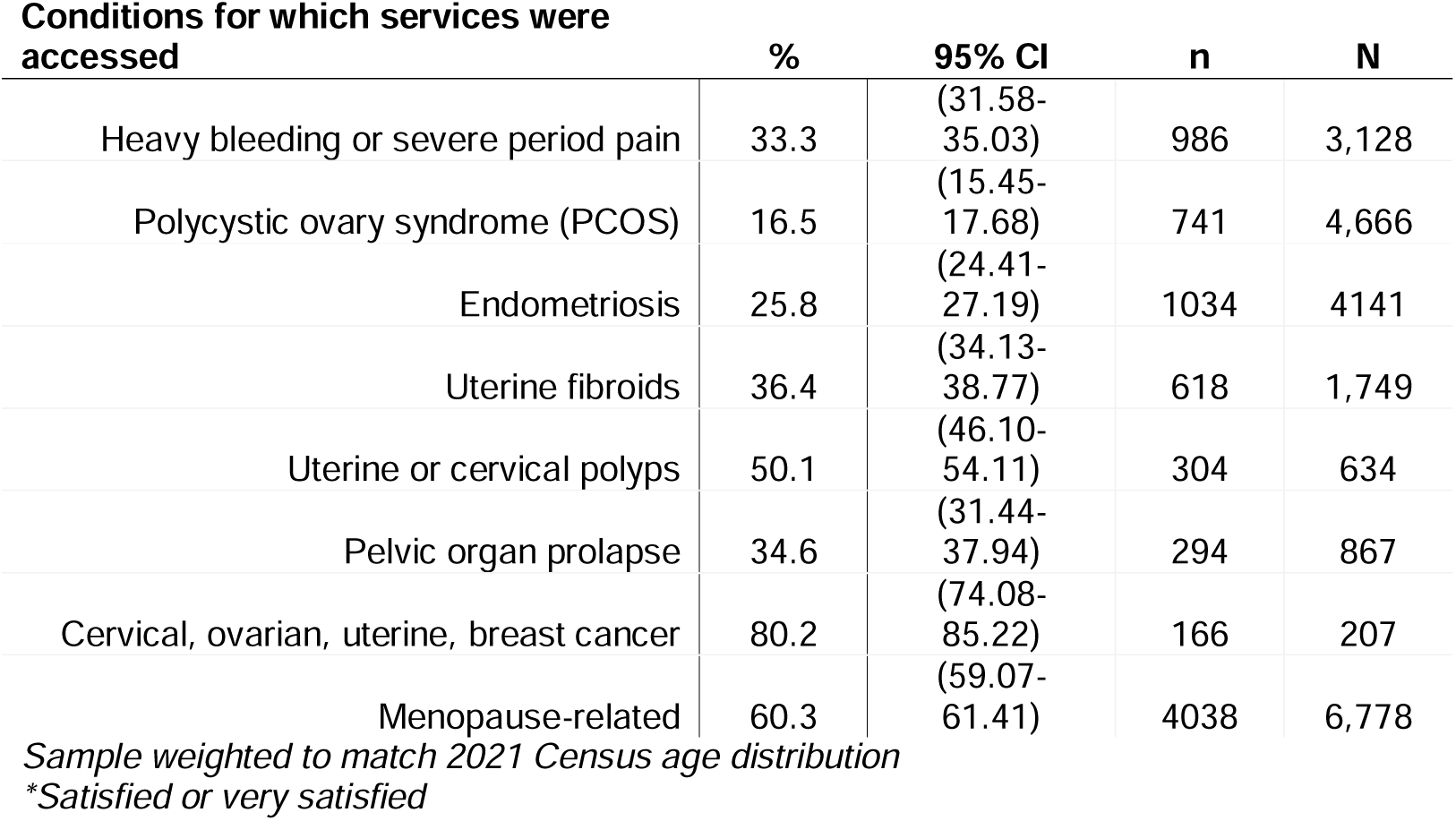
Satisfaction with services*.

|  | % | 95% CI | n | N |
| --- | --- | --- | --- | --- |
| Heavy bleeding or severe period pain | 33.3 | (31.58-35.03) | 986 | 3,128 |
| Polycystic ovary syndrome (PCOS) | 16.5 | (15.45-17.68) | 741 | 4,666 |
| Endometriosis | 25.8 | (24.41-27.19) | 1034 | 4141 |
| Uterine fibroids | 36.4 | (34.13-38.77) | 618 | 1,749 |
| Uterine or cervical polyps | 50.1 | (46.10-54.11) | 304 | 634 |
| Pelvic organ prolapse | 34.6 | (31.44-37.94) | 294 | 867 |
| Cervical, ovarian, uterine, breast cancer | 80.2 | (74.08-85.22) | 166 | 207 |
| Menopause-related | 60.3 | (59.07-61.41) | 4038 | 6,778 |

*Sample weighted to match 2021 Census age distribution*
*\*Satisfied or very satisfied*

## Discussion

### Summary of main results

This is to our knowledge the first study to assess reproductive health service access in England, across three reproductive health domains, based on self-reported symptoms and conditions. Overall, respondents with a reproductive health need had low access to services for their symptoms and conditions, ranging from just over a third for heavy bleeding or severe period pain to just over a half for gynaecological services. Only a third of respondents with heavy bleeding or severe period pain accessed services for their symptoms or conditions. Just under a half of respondents aged 40-55 years accessed help or advice for menopause-related symptoms, with 48% receiving treatment. The percentage of respondents reporting receiving help or advice and receiving treatment was similar. When controlling for age, differences in menopause and gynaecological service access by financial situation and ethnicity were observed. Respondents with a degree had greater access to services for heavy bleeding or severe pain and gynaecological symptoms and conditions. The main differences by region were in access to services for heavy bleeding or severe pain-with access in all regions lower than London. Satisfaction was highest for those accessing health services for cancer-related conditions and lowest for accessing services for PCOS.

### Interpretation

We know a high percentage of women and people assigned female in England live with reproductive health morbidities (3) and we can assume this group requires services to improve and support their reproductive health and well-being. The finding that Black ethnic groups with a need for gynaecological services had a higher percentage of access to services could reflect higher prevalence of fibroids and severity of fibroid-derived symptoms experienced in this group (17, 18). Greater access to services in London is likely due to greater availability of services and transport networks in the city compared to other regions.

This study cannot be interpreted without acknowledging the socioeconomic intersectionality related to reproductive health service access. Multifactorial conditions such as structural racism and economic deprivation compound inequities in educational attainment and can lead to barriers in identifying a health need, locating available services, and navigating the health system to achieve a satisfactory outcome (19, 20). In terms of clinical presentation, research suggests that women from ethnic minorities may present with different perimenopause/menopause symptoms compared to White women; lack of knowledge and awareness of this among some primary care clinicians may lead to incorrect clinical interpretations and action (21).

The similar percentage of respondents receiving help or advice and receiving treatment, may indicate that once services are accessed, there is a high chance of receiving treatment. There will be people who receive treatment but who haven’t received help, for example those with repeat HRT prescriptions.

### Strengths and limitations

The 2023 Reproductive Health Survey for England received almost 60,000 responses in just six weeks, which indicates an eagerness in the population to report on their reproductive health experiences (3). Despite the large sample size, the sample is not statistically representative of the population due to its non-probability online design meaning that it is limited in generalising to the England population. People with lower educational attainment and minority ethnic groups were proportionately under-represented in the survey. Because these demographic groups are less represented in health research (22) and suffer higher levels of reproductive morbidity (23–26), it is reasonable to assume that the low level of access to services estimated in this study is an underestimate of the true level of service access.

Reproductive service need in this study is based on reported symptoms or conditions in each domain. Measures of service access in each domain was reliant upon respondents’ self-reported need. If need was under-reported (or missing), our estimates of service access would be lower than the true estimates if these respondents were less likely to access services. Missing data for the need and access survey items was low (<4%). The observational design means that service access differences by demographic characteristics cannot be attributed to the characteristics itself, i.e. the cause of the differences is not necessarily due to the characteristic, rather to unmeasured or a combination of factors not examined in this study.

While this study provides estimates of service access and indications of variation by demographic characteristics, it cannot provide detailed contextual explanations for the low service access. The study also cannot determine if respondents tried but were not able to access services and why. For example, financial constraints, caregiving responsibilities and restrictive workplace policies can be barriers to healthcare seeking, particularly for women (27). Data on satisfaction with services accessed was by definition, among respondents who managed to access a service. We did not look at what types of services women were accessing or how many they had accessed and how this may have affected satisfaction. Ease in which the appropriate service is identified and in which an appointment can be made can influence how satisfied a person is with the service. Someone could be dissatisfied with a service because they were not able to get it, but this was not captured in the survey. In relation to contraception, for example, it has been observed that type of service accessed varies for user demographic characteristics and that multiple services can be used to obtain supplies (28). We need more research to understand the barriers and to characterise the attempts and pathways to access-be them successful or unsuccessful. This information will help us understand what people need, what they are not able to get and why they were not able to get it.

### Implications for research, policy and practice

England’s National Health Service has set out its 10-year Health Plan (2025–2035) (29), with a focus on embracing digital transformation and preventative healthcare, restructuring NHS governance, and relocating care into communities, for example implementing Family Hubs in all communities. There is, however, a critical absence of specifically highlighting reproductive health, with the words *reproductive, reproduction* or *menopause* not included at all in the 171-page document. The word *women* is mentioned only 17 times: five times in a Tower Hamlets case study box, and twice in a reference.

Our study reports low access to reproductive health services, among those symptoms and conditions, across their reproductive lifecycle. Heavy menstrual bleeding or pain, gynaecological symptoms and conditions and menopause-related symptoms are often severe and can have detrimental effect on well-being and health. The implications from our research are urgent and include ensuring that the reported poor experiences of access and satisfaction are systematically communicated and used to design planned changes in care and services, via public health, commissioning and health services teams at local government level.

## Conclusions

This analysis of the 2023 Reproductive Health Survey for England has demonstrated that access to reproductive health services is low, across three major reproductive health domains and across a range of demographic characteristics. We know that the reproductive health burden in England is high, meaning a large percentage of people in England are going about their daily lives putting up with symptoms and conditions-most of which are preventable and treatable. Urgent action is needed to increase access to high quality reproductive health services.

## Data Availability

All data that support the findings of this study are available from the authors upon reasonable request and with permission of Department of Health and Social Care.

## Abbreviations

SRH: Sexual and reproductive health
RCOG: Royal College of Obstetricians & Gynaecologists
RHSE 2023: 2023 Reproductive Health Survey for England
PCOS: Polycystic ovary syndrome

## Declarations

### Ethics approval and consent to participate

Approval for this study, in accordance with the Declaration of Helsinki, was granted by the London School of Hygiene and Tropical Medicine Ethics Committee (LSHTM Ethics Ref: 29389) on 25th July 2023. After clicking the online survey link, survey respondents were taken to the participant information sheet and consent form, where they provided voluntary informed consent.

### Consent for publication

Not applicable.

### Availability of data and materials

The data that support the findings of this study are available from Department of Health and Social Care but restrictions apply to the availability of these data, which were used under license for the current study, and so are not publicly available. Data are however available from the authors upon reasonable request and with permission of Department of Health and Social Care.

### Competing interests

The authors declare that they have no competing interests.

### Funding

This study is funded by the National Institute for Health and Care Research, Policy Research Programme, NIHR206128. The views expressed are those of the author(s) and not necessarily those of the National Institute for Health and Care Research or the Department of Health and Social Care.

### Author contributions

RSF, MJP, and OLM conceptualised, designed and led the 2023 Reproductive Health Survey for England, including questionnaire development, recruitment strategy and data collection. OLM led the data analysis for this article, with input from MJP and RSF. OLM drafted the manuscript, with contribution from all authors over several iterations. All authors were responsible for reviewing and editing the manuscript. All authors have read and approved the final version for publication.

## Acknowledgements

Not applicable.

## Notes

### Competing Interest Statement

The authors have declared no competing interest.

### Author Declarations

The Ethics Committee of the London School of Hygiene and Tropical Medicine gave ethical approval for this work (LSHTM Ethics Ref: 29389).

